# K-mer based method for finding sequence type-specific PCR primers for *Listeria monocytogenes*

**DOI:** 10.64898/2025.12.01.25341377

**Authors:** Reidar Andreson, Age Brauer, Lauris Kaplinski, Merle Külaots, Signe Saumaa, Ants Kurg, Maido Remm

## Abstract

*Listeria monocytogenes* is a deadly foodborne pathogen for which rapid identification of outbreak strains is critical. Yet current sequence typing methods such as multi locus sequence typing or whole-genome sequencing are time consuming and resource intensive. In this study, we developed a k-mer-based computational approach to pinpoint genomic regions unique to specific *L. monocytogenes* sequence types and designed polymerase chain reaction primers targeting each type’s signature sequences. We screened thousands of genomes to identify robust type-specific markers, then validated the resulting primer sets on 51 isolates representing 17 sequence types in both singleplex and multiplex PCR assays. The primers successfully identified their target sequence types with 100% sensitivity and specificity across the tested isolates, even when assays were combined in multiplex format. This k-mer guided primer design strategy enables rapid and low-cost sequence typing of *L. monocytogenes* for routine screening and outbreak response. It bridges the gap between lengthy genomic sequencing workflows and the need for immediate field ready strain identification in food safety or other public health contexts.

## 1 Introduction

*Listeria monocytogenes* is a facultatively anaerobic Gram-positive foodborne bacterium that is causing fatal disease listeriosis with high mortality rates up to 30% (Mead et al., 1999). Infections affect mostly elderly population or immunocompromised patients, but also pregnant women and newborns (Farber and Peterkin, 1991). *L. monocytogenes* can easily adapt to harsh environments with high salt concentrations (Burall et al., 2015), cold temperatures (Muchaamba et al., 2021), acidity (Cheng et al., 2015) and chemical treatments (Duze et al., 2021). The bacterium uses many genes (roughly seven percent of its genome) for adapting to specific environmental conditions (Glaser et al., 2001). It is also shown that *L. monocytogenes* is naturally resistant to multiple types of antibiotics and therefore rapid finding of the source of outbreaks and identification of pathogenic strains quickly is critical (Olaimat et al., 2018).

Although various alternative methods ranging from serotyping to banding patterns (e.g., RFLPs, AFLPs) have been implemented over the years, currently the most popular method for genotyping major clonal groups and subtypes of *L. monocytogenes* is through the multi locus sequence typing (MLST) scheme (Ragon et al., 2008; Nyarko and Donnelly, 2015). The MLST proof-of-concept method is based on identification of DNA sequences of a small number of hypervariable conserved housekeeping genes and comparing their mutation profiles (Maiden et al., 1998; Salcedo et al., 2003). Ongoing development of next generation sequencing technologies has made it possible to identify full genomes of large number bacterial isolates during foodborne disease outbreaks. Furthermore, gene-by-gene allelic profiling of core genome genes or the core genome MLST (cgMLST) has been developed to become the main diagnostic tool for strain typing (Maiden et al., 2013; Ruppitsch et al., 2015). It is even shown that the discriminatory power of cgMLST clearly exceeds pulsed-field gel electrophoresis (PFGE) typing, which is often considered as the gold standard for subtyping of *L. monocytogenes* (Lüth et al., 2018).

However, there are some major challenges of cgMLST typing due to lack of standardized nomenclature to exchange data between agencies within and between different countries (Moura et al., 2016). During foodborne disease outbreaks it is crucial to timely track the source of bacterial contamination (Jackson et al., 2016). Sending samples into public health agencies to run whole genome sequencing may take weeks to get results. Also, the cost of sequencing and storing data is another criterion that needs to be considered when analyzing large quantities of samples on a regular basis.

In contrast, k-mer based classifiers such as Kraken2 (Wood et al., 2019), StrainScan (Liao et al., 2023) or StrainSeeker (Roosaare et al., 2017) are optimized for taxonomic/strain identification from read k-mers and not for detecting MLST-specific regions from genome. Kraken2 uses exact k-mer matches and lowest-common-ancestor assignment to label reads against a reference database that is fast and sensitive for detection, but it does not output standardized allelic profiles. StrainSeeker similarly maps sample-specific k-mer sets onto a user supplied guide tree to report the best matching strain/clade rather than alleles. StrainScan extends this idea with a tree-based k-mer index to resolve known strains in metagenomic or isolate data. However, it again relies on database encoded strain signatures rather than calling alleles at defined housekeeping loci. Moreover, the performance of k-mer classifiers is tightly coupled to the completeness and composition of their reference databases and can degrade when the sample’s strain diverges from available references. These conditions can complicate their use for generating portable MLST genotype designations.

Recent tools illustrate adjacent ideas, but do not directly solve MLST type-specific primer design. PUPpy (Ghezzi et al., 2024) automates discovery of phylogenetically unique targets within a defined community and then designs microbe- or group-specific primers for strain level detection and absolute quantification. It operates on coding sequences provided by the user and reports specific assays rather than alleles or specific markers tied to MLST loci.

Another tool called varVAMP (Fuchs et al., 2025) focuses on highly variable viral genomes, using an MSA-derived consensus and a k-mer-based enumeration around Primer3 (Kõressaar et al., 2018) to create degenerate and tiled primer schemes for full-genome sequencing and qPCR. However, it is aimed at pan-specific coverage across variants, not discrete type-specific diagnostics. SURE-Pipe^1^ focuses on shared/unique region extraction by orchestrating BLAST/BEDTools (Quinlan and Hall, 2010) within a Nextflow workflow to report regions unique to a target versus non-targets. It’s general purpose is for genome comparison and does not natively couple uniqueness calls to type-specific nomenclature or downstream multiplex primer selection.

In this work, we introduce a k-mer based approach to identify MLST type-specific regions from bacterial whole genomes and create unique PCR primers for each type. We have used 51 *L. monocytogenes* samples covering 17 different sequence types to experimentally test the success rate of given primers in singleplex and multiplex form.

## 2 Materials and methods

### 2.1 Bacterial reads and isolates

For identifying type-specific regions of *L. monocytogenes* we needed genomic reads from the ENA database (O’Cathail et al., 2025). In total, 28545 isolate samples with Illumina paired-end reads were downloaded. These samples included raw sequencing data from various projects around the world. For experimental validation, we had DNA samples from 51 bacterial isolates of 17 different MLST types (designated ST) (ST5, ST7, ST8, ST9, ST29, ST37, ST87, ST101, ST121, ST155, ST173, ST177, ST425, ST451, ST551, ST580, ST1247). For each type, there was DNA of three isolates, a total of 51 samples (Table 1). DNA samples of isolates were obtained from The National Centre for Laboratory Research and Risk Assessment (LABRIS, Estonia), where the MLST types of these samples had been previously determined by whole genome sequencing data analysis. All these *L. monocytogenes* samples were collected from Estonia. Among them were ST1247 isolates, which received widespread resonance in local and foreign press (Mäesaar et al., 2021).

**Table 1.** DNA isolates used for the validation and indications of their types.

| Isolate ID | MLST type | Clonal complex | Isolate ID | MLST type | Clonal complex |
| --- | --- | --- | --- | --- | --- |
| 1 | ST87 | CC87 | 25 | ST551 | CC8 |
| 2 | ST87 | CC87 | 26 | ST551 | CC8 |
| 3 | ST87 | CC87 | 27 | ST551 | CC8 |
| 4 | ST8 | CC8 | 28 | ST580 | CC9 |
| 5 | ST8 | CC8 | 29 | ST580 | CC9 |
| 6 | ST8 | CC8 | 30 | ST580 | CC9 |
| 7 | ST9 | CC9 | 31 | ST177 | CC177 |
| 8 | ST9 | CC9 | 32 | ST177 | CC177 |
| 9 | ST9 | CC9 | 33 | ST177 | CC177 |
| 10 | ST37 | CC37 | 34 | ST451 | CC11 |
| 11 | ST37 | CC37 | 35 | ST451 | CC11 |
| 12 | ST37 | CC37 | 36 | ST451 | CC11 |
| 13 | ST155 | CC155 | 37 | ST173 | CC19 |
| 14 | ST155 | CC155 | 38 | ST173 | CC19 |
| 15 | ST155 | CC155 | 39 | ST173 | CC19 |
| 16 | ST101 | CC101 | 40 | ST425 | CC90 |
| 17 | ST101 | CC101 | 41 | ST425 | CC90 |
| 18 | ST101 | CC101 | 42 | ST425 | CC90 |
| 19 | ST121 | CC121 | 43 | ST7 | CC7 |
| 20 | ST121 | CC121 | 44 | ST7 | CC7 |
| 21 | ST121 | CC121 | 45 | ST7 | CC7 |
| 22 | ST5 | CC5 | 46 | ST29 | CC29 |
| 23 | ST5 | CC5 | 47 | ST29 | CC29 |
| 24 | ST5 | CC5 | 48 | ST29 | CC29 |
|  |  |  | 49 | ST1247 | CC8 |
|  |  |  | 50 | ST1247 | CC8 |
|  |  |  | 51 | ST1247 | CC8 |

### 2.2 Genome assembly and determination of MLST types

To determine the MLST types of downloaded public strains, we used the computational workflow described below. First, the reads were cleaned of low quality nucleotides, and adapter sequences were removed. For this, the *fastp* program (Chen et al., 2018) was used with the following parameters: minimum read length 50 nucleotides (*--length_required 50*), average quality 30 (*-cut_mean_quality 30*), *--cut_tail, --cut_front*, and *--detect_adapter_for_pe*. In the next step, the cleaned reads were assembled with *SPAdes* (Prjibelski et al., 2020) into genomic contig sequences using the *--isolate* parameter. Samples with the longest contig of <50,000 nucleotides and an average sequencing depth of less than 20 were filtered out from the results. The final step was the determination of the MLST type by the MLST^2^ program, for which the default parameters and the PubMLST (Jolley et al., 2018) dataset containing >2300 different MLST types of *L. monocytogenes* was used.

### 2.3 Phylogenetic tree of MLST types

A prerequisite for the construction of a tree is the creation of distance matrix based on sequence comparisons. For this, we used the *dashing* program (Baker and Langmead, 2019), that created a distance matrix based on sequencing reads. When running the program, the following parameters were changed: the length of the k-mers 32 nucleotides (*-k 32*), *--sketch-size 20*, -*-use-range-minhash*, and *--full-mash-dist*. From the created distance matrix, a phylogenetic tree in newick format was compiled using the *quicktree* (Howe et al., 2002) program (with default parameters). The results were visualized with the MEGA11 program (Tamura et al., 2021).

### 2.4 Search for type-specific k-mers

As a first step, it was necessary to create a list of k-mers for each sample of ENA reads. To do this, we used the GenomeTester4 package (Kaplinski et al., 2015) program *glistmaker*, for which we set the length of the k-mer to 32 nucleotides (*--wordlength 32*). Previous work by our group has shown that 32 could be optimal for finding more specific k-mers (Raime and Remm, 2018).

To create a common list of non-targets, the GenomeTester4 package program *glistcompare* was used with parameters *--union* and a frequency limit value of 5 (*--cutoff 5*) for k-mers. The latter is necessary to exclude very rare (<5 times) k-mers, which are most likely sequencing errors.

Similarly to the above, using *glistcompare* a list of k-mers was created with the parameters *--intersection* and the frequency limit value of k-mers 5 (*--cutoff 5*). Those k-mers that were not represented in all samples were removed. This was necessary to eliminate k-mers, which are present only in a particular sample and are therefore not characteristic of all members of a given type.

Next, we compared the common list of each type with the combined list of non-targets to exclude k-mers that are also present in types other than the target. To do this, we used the *glistcompare* with the parameter *--difference*.

### 2.5 Design of type-specific PCR primers

Type-specific k-mers were mapped to their genomic coordinates using NCBI BLAST (Altschul, 1997). The package program *blastn* was used with an alignment percentage of 100 (*-perc_identity 100*) and a query sequence coverage percentage of 100 (*-qcov_hsp_perc 100*). The resulting HSP coordinates for each k-mer were recorded for downstream processing.

K-mers whose mapped coordinates overlapped or were directly adjacent on the same contig/strand were merged into contiguous genomic intervals as type-specific regions. Candidate intervals were restricted to sizes compatible with PCR using a product length window of 60–1000 nt. Intervals shorter than 60 nt (e.g. single isolated k-mer hits) and >1000 nt were excluded from primer design.

For every retained region, PCR primers were designed with Primer3 using following settings: primer length: min 18, opt 20, max 22 nt; product size range: 60–1000 nt; maximum allowed difference between primer pair melting temperatures (Tm): 3 ^°^C. All other Primer3 parameters were left at their defaults.

To assess potential cross-reactivity, all candidate primers were aligned to the non-target genome set using *blastn*. In this step, we permitted sequence mismatches, but required full-length primer coverage: -*qcov_hsp_perc 100* (no *-perc_identity* constraint). All HSPs and their genomic coordinates were retained for pairwise evaluation. A pair was retained if: (a) at least one primer had no non-target hit, or (b) both primers hit but not in opposing orientations 1–1000 nt apart (no plausible amplicon), or (c) both primers hit at 1–1000 nt spacing but at least one had a mismatch within the 3′ terminal 6 nt. Only pairs meeting more than one of these conditions was kept.

### 2.6 DNA extraction

The genomic DNA of *L. monocytogenes* isolates were purified with GeneJET Genomic DNA purification Kit (Thermo Scientific) by using “Gram-positive Bacteria genomic DNA Purification Protocol” according to manufacturer’s protocol, except for the first step. As isolates of *L. monocytogenes* were obtained growing on blood agar, the bacterial cells were first collected from agar by using an inoculation loop and after that were suspended in Gram-positive bacteria lysis buffer. All procedures were done in strict rules and conditions of Biosafety Lab 2 in Institute of Molecular and Cell Biology, University of Tartu.

### 2.7 PCR experiments

To check the specific PCR primers designed, we conducted laboratory experiments. We had the possibility to use DNA samples from 17 different types of isolates and each type in three replicates. In total, it had formed 51 singleplex reactions in which only one pair of primers is used at a time. Additionally, the success rate of universal primers was controlled under the same reaction conditions. Ideally, only a single product (band on the gel) should arise with primers of the corresponding type and other types should not show any yield in given reaction. Universal primer pair should ideally work with all samples.

#### Singleplex solution and program

10 μl reaction volume:10x buffer (Solis Biodyne) 1μl; 25 mM MgCl_2_ (Solis Biodyne) 0,6 μl; 2.5 mM dNTP mix (Solis Biodyne) 1.2 μl; Hot FirePol (Solis Biodyne) (5U/μl) 0.1 μl; MQ water 5,1 μl; Primers (10nM (5 F+5 R))1 μl; DNA (5ng/μl) 1 μl. PCR program: 95°C 15 min; 95°C 30 s, 60°C 30 s - 30x,72°C 40 s; 72°C 10 min.

In multiplex PCR experiments, the desired number of primer pairs are mixed in one reaction, which allows saving the costs and time of the reagents. As a result, the gel image should develop several bands of the correct length instead of one correct band. Since in some cases bands of several products are expected to be performed on the track of a single gel because of a multiplex test, it is extremely important that the lengths of the products are different enough to achieve visual resolution. Therefore, we have checked with the MultiPLX (Kaplinski et al., 2005) program (parameters -cutoff1 −3.0 -cutoff2 −10 -cutoff3 −13.0) that primer pairs do not create strong secondary structures among themselves, which can negatively affect the success of PCR, and are divided into groups with separable product sizes.

#### Multiplex solution and program

10 μl reaction volume: 10x buffer (Solis Biodyne) 1μl; 25 mM MgCl_2_ (Solis Biodyne) 0,6 μl; 2.5 mM dNTP mix (Solis Biodyne) 1.2 μl; Hot FirePol (Solis Biodyne) (5U/μl) 0,1 μl; MQ water 4.5 μl; Primers (pairs mixed together) (10nM) 1.5μl; Primer Univ_6 (10nM) 0,1 μl; DNA (5ng/μl) 1 μl. PCR program: 95°C 15 min; 95°C 30 s, 58°C 30 s - 31x, 72°C 40 s; 72°C 10 min.

## 3 Results

### 3.1 Identifying type-specific k-mers from genomes

In order To find unique type-specific k-mers for *L. monocytogenes* samples, we downloaded 28,545 public ENA Illumina paired-end read sets. These were quality filtered and adapter trimmed, resulting high quality reads suitable for the assembly. Genome assemblies were generated with SPAdes and screened. Datasets with insufficient contiguity or depth were excluded from downstream analysis. For the retained samples, sequence types were successfully assigned using the pubMLST scheme for *L. monocytogenes*. The MLST type remained undetermined for 980 out of 28,545 and 1252 different types were determined in total. Samples for which the species were determined to be something else than *L. monocytogenes* were removed from the dataset. The final number of samples remaining was 27,206 (Supplementary Table S1).

Next, we decided to construct a phylogenetic tree of isolates based on reads to see if the isolates with the same MLST type cluster together in a clade or are more broadly distributed. In the latter case, it may be more difficult to find type-specific regions as genomes share common regions with many different types. Since with all 27,206 samples this data analysis would be too time consuming and difficult to visualize later, we decided to take ten random samples for each type (out of 1252), which occurred at least 10 times in the dataset. In addition, we checked that all 17 types of our experimental dataset were included. In total, 1168 samples of various types were used to build a phylogenetic tree.

The phylogenetic tree shows the distribution of sequence types along the branches, with the 17 sequence types of interest highlighted in different colors (Supplementary Figure S1). Fifteen of these 17 types cluster relatively compactly. The exceptions are ST37 and ST173, which are distributed across several major branches. ST9 and ST580 form a distinct group, and a similar pattern is observed for ST8, ST551 and ST1247. In this case, such groupings are consistent with their presence in the corresponding clonal complexes CC9 and CC8. A clonal complex is defined as a set of sequence types grouped according to the inferred evolutionary descent of isolates under the assumption of a shared common ancestor. The name of the complex typically reflects the putative ancestral type (e.g. ST8 for CC8). Overall, this grouping pattern suggests that it should be feasible to identify type-specific genomic regions and to design corresponding PCR primers.

From 27,206 ENA read sets, we enumerated 32-nt k-mers for each sample and used these to assemble two complementary resources: a comprehensive non-target background and type-specific core repertoires. Figure 1 shows the structure of the workflow, by which it was possible to create lists of k-mers of samples from targets and non-targets. One of the types was chosen as the target and the rest remained among the non-targets.

**Figure 1.**
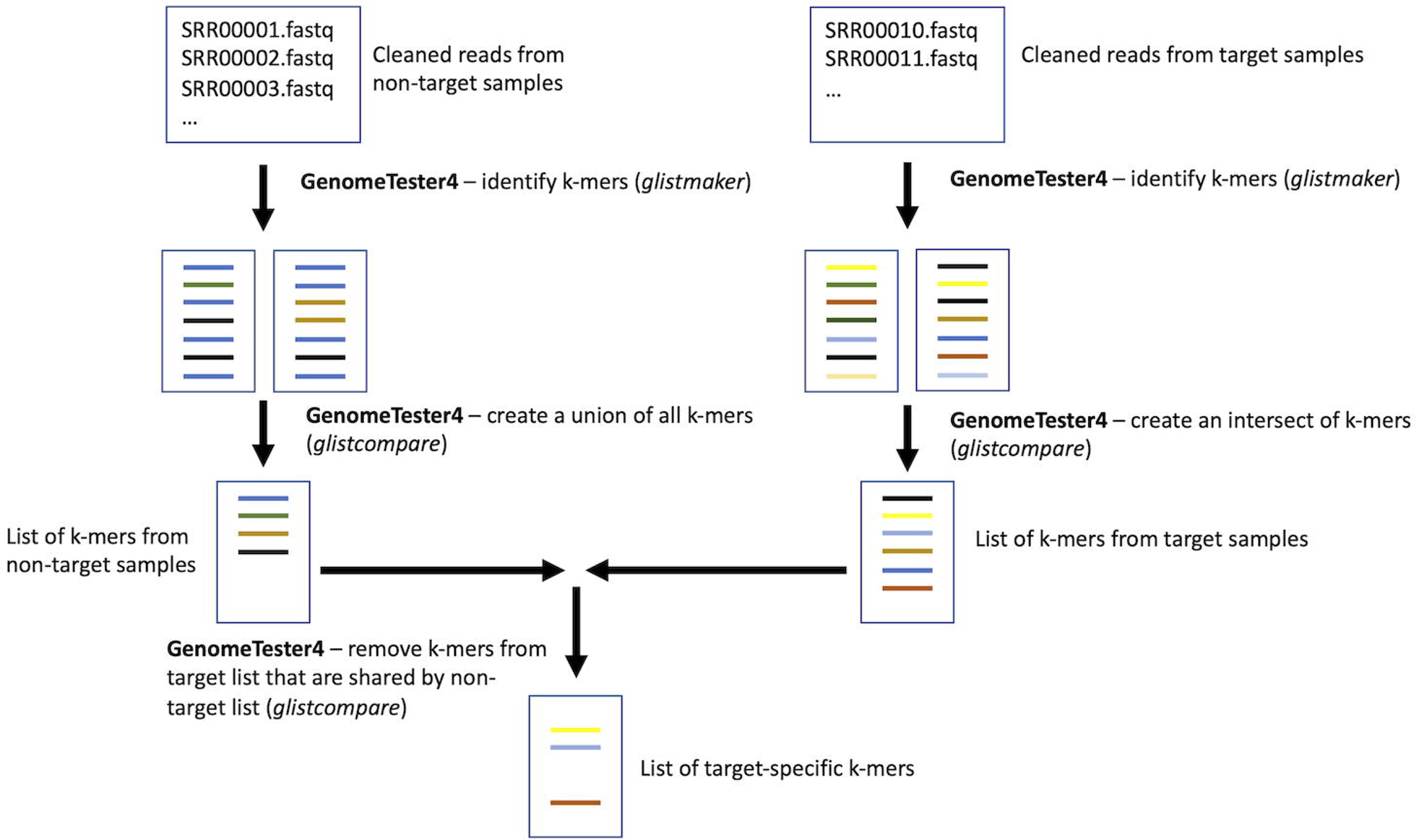
The flowchart of identifying type-specific k-mers.

Aggregating all non-target samples produced a unified background of k-mers while filtering out very rare instances (occurring <5 times), thereby minimizing the influence of sequencing errors. Secondly, for each of the 17 target types, we derived a stringent “core” set by retaining only those k-mers observed (≥5 instances) in every sample of that type, which removed k-mers confined to individual specimens and not representative of the given sequence type. Finally, subtracting the non-target background from each type’s core yielded target-specific k-mer panels. These k-mers are absent from all non-target samples yet ubiquitous across the corresponding target type. Given workflow produced high confidence type defining marker sets suitable for downstream detection and classification.

### 3.2 Merging k-mers to type-specific regions and PCR primer design

To find type-specific genome regions for which PCR primers could be designed, specific k-mers were mapped on genome sequence and the corresponding coordinates of hits were recorded. The PCR primer design workflow is graphically described in Figure 2.

**Figure 2.**
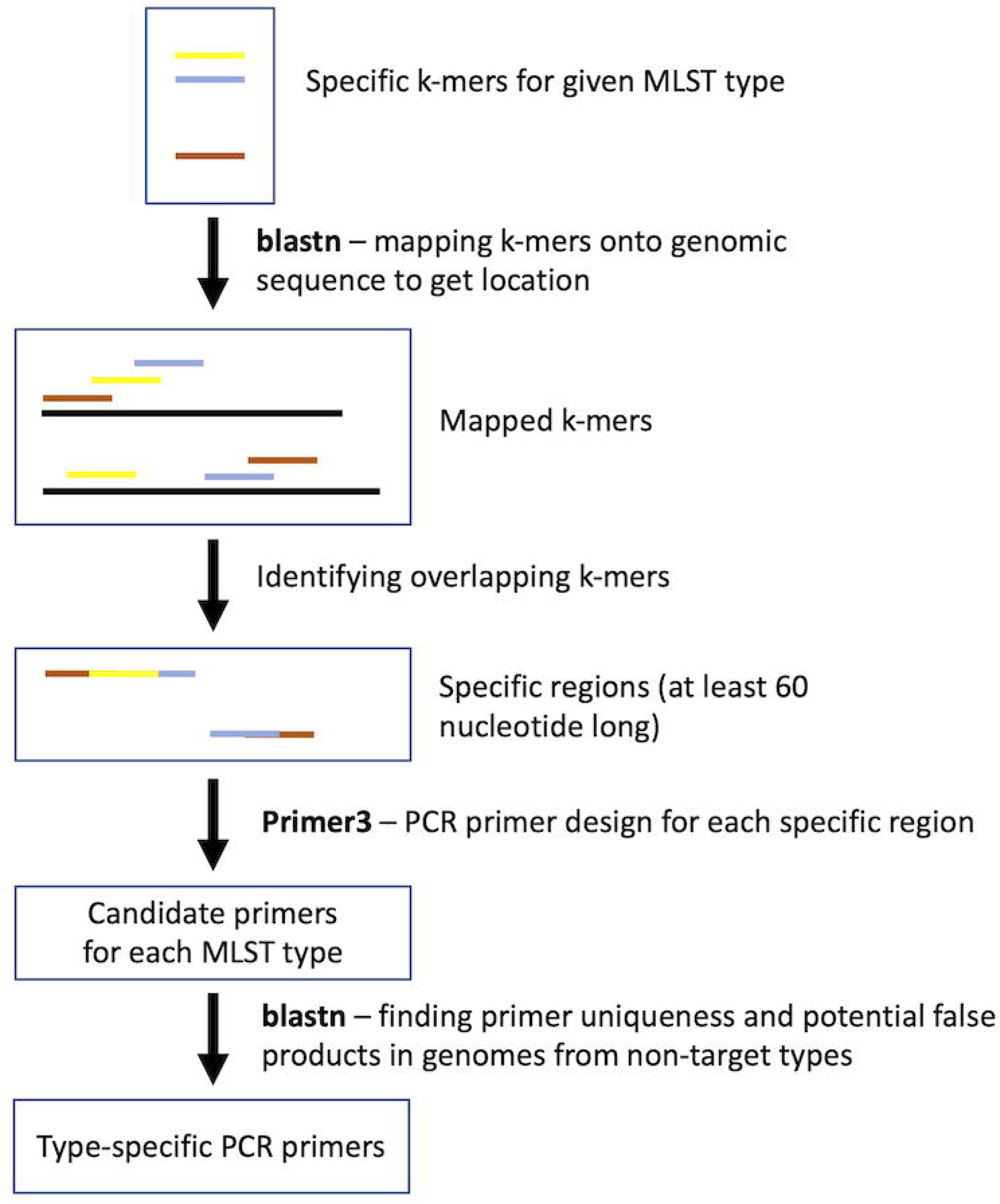
The flowchart of the design of *L*. *monocytogenes* type-specific PCR primers.

The k-mers with genomic positions in the overlay were merged into longer regions based on coordinates. Considering the recommended minimum (roughly two primer lengths) and maximum (<1000 nucleotide) product lengths for the PCR methodology, we set the range to 60-1000. Shorter regions or single k-mers were removed from the selection. As a result, the median value of specific regions across all types was 551.

For each specific region, we tried to design PCR primers with Primer3. Allowing for near identical matches, we realigned all type-specific primers against non-target genomes using full-length (100%) coverage. This secondary screen flagged a subset of primers with potential cross reactivity due to single nucleotide differences, while the remainder showed no full-length hits in non-targets. Primers with such near perfect alignments were marked for exclusion or further scrutiny, and the non-flagged set was carried forward as the high specificity candidate pool.

The result files were analysed and the final primer pairs had to meet the following conditions. The pair was named suitable, (a) if at least one primer mate did not give alignments to any non-target genome of the type; (b) if both primers in the pair did provide alignment from a non-target type’s genome, but not a distance between 1-1000 nucleotides; (c) if both primers in the pair did provide alignment from a non-target type’s genome and a distance between 1 and 1000 nucleotides, but the difference (mutation) lies within the last six nucleotides at the 3’ end of at least one primer.

When we used the full list of samples from ENA database (27,206), problems occurred with some of the target types as very few specific k-mers remained. This may be due to contaminated data, where most of the reads in one sample are indeed from cells of one MLST type, but to a lesser extent from another type. Since our methodology detects all k-mers present in the reads, samples with contaminated reads can end up as false positives among non-targets or targets. In addition, there is always the possibility that sequence type contamination will occur in the process of sequencing, when several samples of *L. monocytogenes* are analysed in the same laboratory.

Therefore, we decided to reduce the amount of data and selected up to 10 random samples (with a total number of 155) from each target type from all ENA data and reran the entire previous workflow with them. This approach made sense, because we had to use only the DNA of these 17 types for experimental control of type-specificity. For types ST173, ST580 and ST1247, we had to use a smaller number of samples than ten, since there were simply no more data in the database. As a result of the analysis, we managed to find type-specific k-mers with a median value of 47000 for 17 ST-s and two CC-s (Table 2). The list of primer sequences and their properties are shown in Supplementary Table S2.

**Table 2:** *L. monocytogenes* type-specific sequences.

| MLST type | CC type | # of ENA samples | # of randomly selected samples | # of type-specific k-mers | # of selected specific regions (>60 nt) | # of PCR primer pairs (product size range 60-1000 nt) | # of PCR primer pairs after filtering |
| --- | --- | --- | --- | --- | --- | --- | --- |
| ST5 | CC5 | 3119 | 10 | 161122 | 1928 | 984 | 17 |
| ST7 | CC7 | 1431 | 10 | 64594 | 687 | 307 | 13 |
| ST8 | CC8 | 569 | 10 | 153 | 1 | 0 | 0 |
| ST9 | CC9 | 1445 | 10 | 47 | 0 | 0 | 0 |
| ST29 | CC29 | 252 | 10 | 39010 | 440 | 206 | 12 |
| ST37 | CC37 | 269 | 10 | 46426 | 444 | 202 | 12 |
| ST87 | CC87 | 345 | 10 | 103565 | 1302 | 704 | 12 |
| ST101 | CC101 | 187 | 10 | 32327 | 429 | 195 | 11 |
| ST121 | CC121 | 623 | 10 | 100178 | 828 | 492 | 12 |
| ST155 | CC155 | 1367 | 10 | 85948 | 627 | 296 | 14 |
| ST173 | CC19 | 5 | 5 | 107450 | 689 | 319 | 14 |
| ST177 | CC177 | 13 | 10 | 57056 | 574 | 262 | 17 |
| ST425 | CC90 | 24 | 10 | 41904 | 444 | 201 | 12 |
| ST451 | CC11 | 163 | 10 | 52729 | 694 | 315 | 11 |
| ST551 | CC8 | 35 | 10 | 789 | 18 | 3 | 0 |
| ST580 | CC9 | 2 | 2 | 1694 | 24 | 6 | 0 |
| ST1247 | CC8 | 8 | 8 | 618 | 11 | 5 | 5 |
| CC8* | CC8 | 612 | 28 | 65936 | 581 | 253 | 10 |
| CC9* | CC9 | 1447 | 12 | 47007 | 533 | 214 | 13 |

\*MLST types marked with coloured background, for which no suitable PCR primer pairs were found, were assembled into one group of clonal complexes named CC8 and CC9. For ST1247, separate primers, specific to this type, were also tested.

### 3.3 Singleplex PCR experiments

The results of the singleplex PCR tests can be seen in Figure 3. A total of 83 different primer pairs were tested on 51 different DNA samples. Three pairs were universal primers (Univ_3, Univ_6, Univ_7), 14 pairs were clonal-complex-specific primer pairs, and the remaining 66 were type-specific. The green boxes represent the correct result (a decent band on the gel) that was supposed to come out (true positive). Yellow colours indicate a false positive result, and white indicates the absence of the expected reaction. For types, there must be 3 green boxes for each pair of primers, for universal ones all must be green, and for clonal complex-specific primers, the types belonging to the corresponding complex must be green (9 DNA for CC8, 6 for CC9). For each type and clonal complex, a smaller number of primers were first tested and if pairs with 100% specificity and sensitivity were not detected, new primers were picked from the list of candidate primers for testing. A total of 4233 PCR experiments were carried out, of which 2813 (∼67%) were successful, i.e. gave the expected result (true positive).

**Figure 3.**
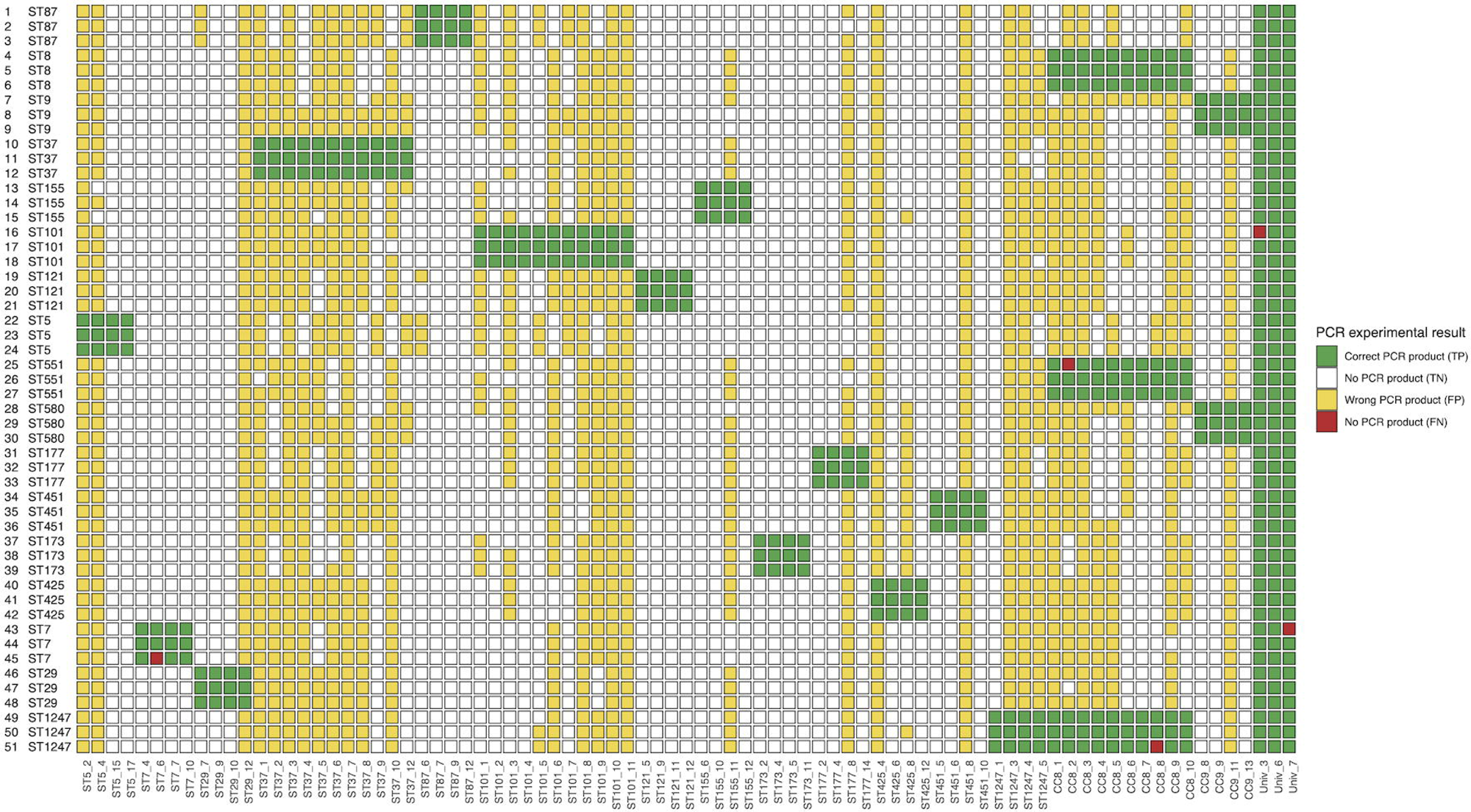
Singleplex PCR experiments. Two columns in the Y axis describe the isolate number and given MLST type accordingly. In X axis, “ST” describes type-specific primer pair and “CC” clonal complex specific primer pair. “Univ” states for universal primer pair that must give a positive result for all samples. TP – true positive, TN – true negative, FP – false positive, FN – false negative.

Closer look at the specificity and sensitivity of primers (Supplementary Table S3) shows that corresponding median values for all types are 89% and 100%. Specificity indicates the ability of the primer pair to distinguish a particular type from the rest, that is the primer pair must not give a PCR product on any other type. Sensitivity indicates how well the primer pair identified the correct type. Only five pairs of primers did not develop the correct positive PCR product with the DNA of all three corresponding types. In 55% of types, specificity was lower than 100%, for which false positive products with DNA of other types were formed.

Based on 10 random ENA samples, it was not possible to find a primer pair with 100% specificity for the ST37 type. For CC8, the best primer pair had 98% specificity and 100% sensitivity (CC8_7). In all other cases we found at least one perfectly working PCR primer pair. From the tested universal primers, the one called Univ_6 had 100% experimental success rate for both specificity and sensitivity.

We also wanted to analyse genomic regions for all designed PCR primers to see, if there are some shared properties. For that we used *blastx* with *L. monocytogenes* reference strain EGD-e genome and protein sequences. Only 11% of the blast hits corresponded to coding regions with known or annotated functions, whereas about 89% were in proteins annotated as hypothetical, putative, or with no characterized function. The hits clustering in surface proteins and regulatory elements may point to repetitive DNA elements in those proteins rather than in unique, highly conserved housekeeping genes. Additionally, a search of the primer alignment coordinates and identifiers revealed no hits within or near MLST (*abcZ*, *bglA*, *cat*, *dapE*, *dat*, *ldh, lhkA*) loci.

### 3.4 5-plex PCR experiments

As the first option for multiplexing, we tried to match a maximum of 5-plex pairs (4-5 pairs of primers in one reaction). To do this, based on the results of previous singleplex experiments, we selected primer pairs whose product lengths would be sufficiently distinguishable from each other (Figure 4A). As a result, we formed three different groups of primers, with five pairs of primers in two and four in one. For the reaction, mixtures of 4 or 5 pairs of primers were mixed and experiments were carried out on the DNA of all 51 isolates.

**Figure 4.**
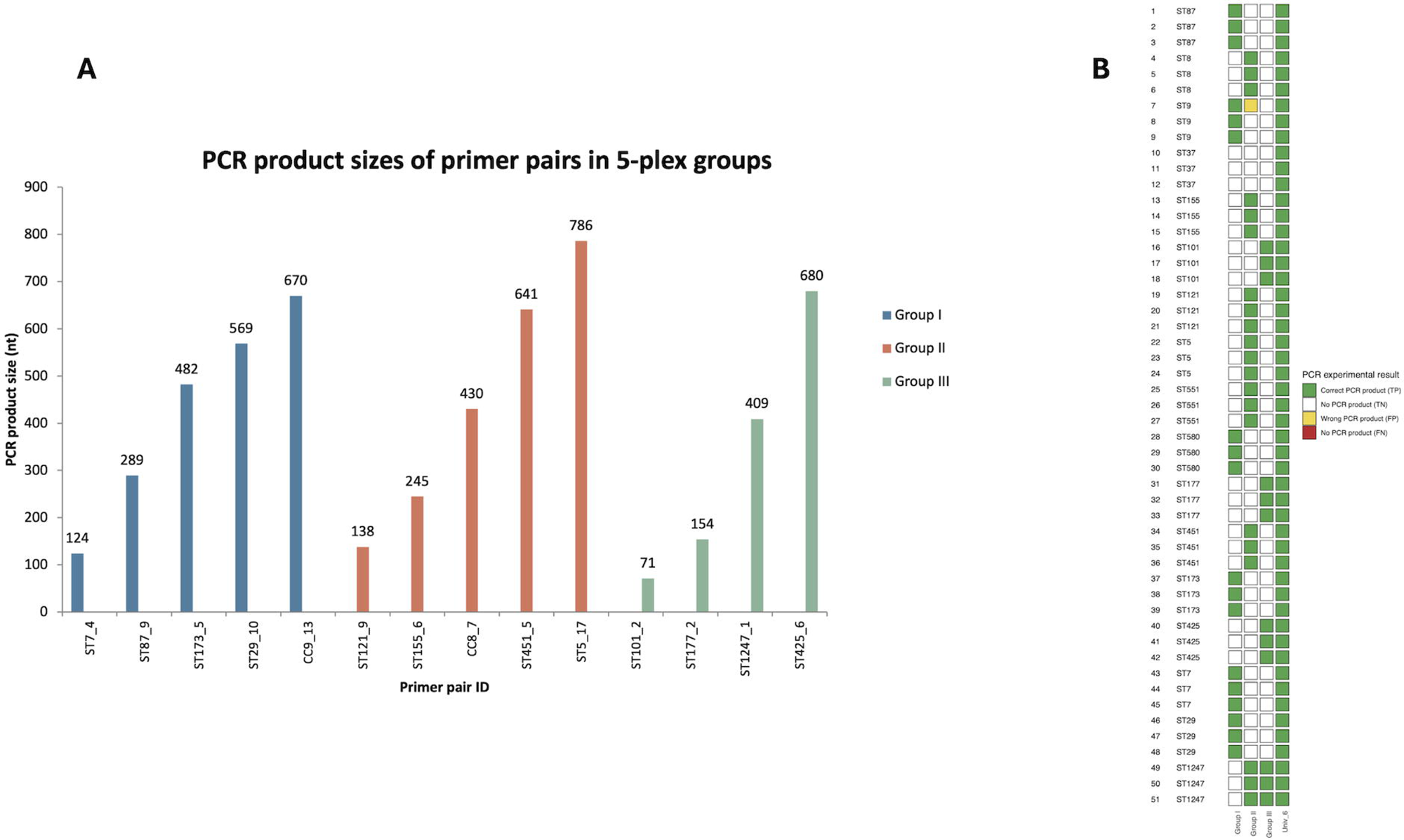
5-plex PCR experiments. In section A, three different multiplex groups were formed that contain primer pairs with product sizes ranging from 71 to 680 nucleotides. In section B, two columns in the Y axis describe the isolate number and given MLST type accordingly. In X axis, “Group I-III” describes three different PCR primer pair groups. “Univ” states for universal primer pair that must give a positive result for all samples. TP – true positive, TN – true negative, FP – false positive, FN – false negative.

The results of the 5-plex multiplex PCR experiment can be seen in Figure 4B. The first group (Group I) gives correct results with six types (CC9 gives with two different types), the second group (Group II) gives with seven types (CC8 with three different ones), and the third group (Group III) with four types. The sensitivity of these groups is 100%, all prescribed types are correctly recognized. In the case of Group II, there was a problem with single isolate (yellow box), which is why the specificity with this group is 98%. Such a result was expected, since it was seen from the singleplex experiments that CC8_7 primer pair gives a nonspecific reaction with the 7th isolate. Summing up, it can be said that the 5-plex experiments were as successful as the singleplex experiments.

### 3.5 15-plex PCR experiment

Based on the success of the previous 5-plex experiments, we decided to take it a step further and try the multiplex PCR reaction with all 14 pairs of primers. Accordingly, we had to slightly change the list of suitable primers, where we replaced the CC8_7 primer pair with CC8_6 variant, since the length of a given product (131 nucleotides) was more easily distinguishable from ST1247 (belongs to the CC8 clonal complex). We mixed 14 primer pairs with the best working universal primer pair (Univ_6) and checked whether PCR products are formed for all 51 isolates and whether they are also of the correct length (Supplementary Table S4).

The result of the 15-plex multiplex PCR experiment can be seen in the gel image in Figure 5. In the central part of the gel is located a line of straight bands with a universal primer. In the case of CC8, there are three bands on tracks 49-51, where one is a product of a universal primer pair with a length of 430 nt, above it a product of the ST1247 type with a length of 409 nt and underneath a product of the CC8 clonal complex with a length of 131. On all other tracks a band of both a universal product and a type-specific product can be seen.

**Figure 5.**
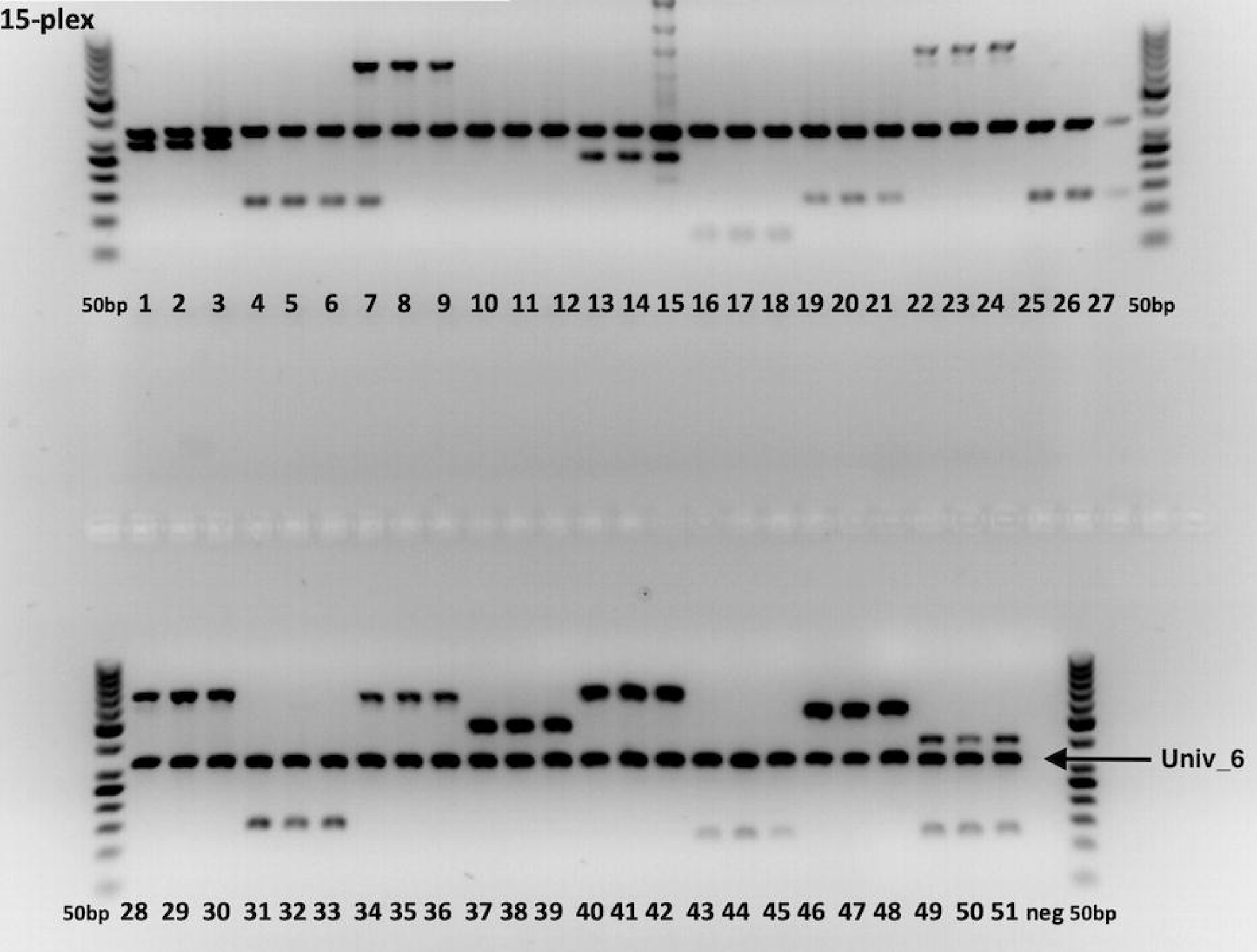
15-plex PCR experimental results. 51 sample DNA-s were screened with 15-plex primer pairs. Single continuous band in the represents the product from the universal primer pair (Univ_6) as a positive control of *L. monocytogenes’* DNA presence. 50bp states as a DNA step ladder.

## 4 Discussion

This study presents and experimentally validates a k-mer driven workflow for discovering MLST type-specific genomic regions in *L. monocytogenes* and designing PCR assays that function in both singleplex and multiplex formats. Using 27,206 public WGS datasets to define type-specific k-mers and 51 locally sourced isolates across 17 MLST sequence types for laboratory validation, we achieved robust type discrimination with median sensitivity of 100% and median specificity of 89% for candidate primer pairs, with several perfect performers per type or clonal complex. These results demonstrate that allele independent k-mer set operations can yield portable, interpretable, and laboratory ready screening markers even when closely related types share substantial genome content.

Our algorithm is distinctive in three ways. First, it uses strict set logic on k-mer inventories like intersection within type and union of non-targets to isolate regions that are both ubiquitous in the type and absent from others and these properties translate directly into reliable PCR targets. Second, it places candidate k-mers onto assemblies with exact match constraints to form contiguous primer sized intervals avoiding fragile point mutation signatures. Third, it demonstrates multiplex readiness by incorporating primer thermodynamics and cross interaction checks from the outset. The result is a pipeline that transforms MLST labels into deployable type-specific assays without requiring whole-genome alignment or special SNP modelling.

For routine screening in food production environments and public health triage, turnaround time and per-sample cost are decisive (Tran et al., 2023). Centralized WGS workflows typically require ∼5–10 days from sample to report when library prep, batching, sequencing, and analysis are considered (Beukers et al., 2021). This can be often longer in public health institutions as their pipelines are optimized for cost over speed. In contrast, PCR-based assays routinely deliver results within 12–24 hours, can be automated at scale, and are far cheaper per sample. These enable high-throughput and rapid decision making in line clearance or targeted sanitation, for example. Thus, the development and usage of multiplex PCR panels offers a pragmatic balance between speed, cost, and epidemiologic resolution.

Our approach depends on the representativeness and quality of public data used to identify type-specific regions. While our curation mitigated some contamination and imbalance, continued updates to the non-target set and routine rescreening of candidate regions will be necessary as new diversity is deposited. PCR experiment performance can also be affected by DNA source (food vs clinical), DNA quality and thermocycler conditions. The additional *in silico* checks with an updated screenings against expanding *L. monocytogenes* genomes and optimizations of wet-lab protocol can further reduce some sporadic false positive results. Although we have validated 5-plex and 15-plex formats with high success, higher-plex panels may require some pipeline or protocol redesign to preserve clear gel separation or transition to qPCR readouts.

## 5 Conclusion

We have linked *L. monocytogenes* MLST labels to k-mer defined type-specific regions and thus created a practical route from population nomenclature to rapid type identification. The singleplex and multiplex results across 17 MLST types indicate that this strategy is viable for front line industrial screening and outbreak triage. Although periodic off-target rescreening is advisable as databases grow, given approach yields portable, interpretable, and multiplex PCR assays that performed well across diverse list of MLST types. Given approach can be easily extended to additional *Listeria* lineages and other bacterial taxa.

## Data availability statement

The ENA identifiers used in this study can be found in Supplementary Table S1. All custom scripts used in this work are available at https://github.com/bioinfo-ut/MLST-specific_genomic_regions.

## Author contributions

RA: Investigation, Software, Methodology, Validation, Visualization, Writing – original draft. AB: Conceptualization, Methodology, Writing – review & editing. LK: Conceptualization, Investigation, Writing – review & editing. MK: Methodology, Visualization, Writing – review & editing. SS: Methodology, Validation, Writing – review & editing. AK: Conceptualization, Investigation, Writing – review & editing. MR: Conceptualization, Investigation, Funding acquisition, Project administration, Supervision, Writing – review & editing.

## Acknowledgements

We thank Toomas Kramarenko from LABRIS for providing the isolates of *L.monocytogenes*. Some parts of the data analysis was performed using the facilities of the High-Performance Computing Center at the University of Tartu (https://doi.org/10.23673/PH6N-0144).

## Footnotes

1 https://github.com/BPaul-bioinfoLAB/SURE-Pipe

2 https://github.com/tseemann/mlst

